# Advancing 90-day mortality and anastomotic leakage predictions after oesophagectomy for cancer using explainable AI (XAI)

**DOI:** 10.1101/2024.04.09.24305451

**Authors:** Sebastian Djerf, Oscar Åkesson, Magnus Nilsson, Mats Lindblad, Jakob Hedberg, Jan Johansson, Attila Frigyesi

**Author notes:** Equal authors (Jan Johansson, Attila Frigyesi).

## Abstract

Oesophagectomy for cancer of the oesophagus carries significant morbidity and mortality. Ninety-day mortality and anastomosis leakage are critical early postoperative problems traditionally analysed through logistic regression. In this study, we challenge traditional logistic regression models to predict results with new explainable AI (XAI) models. We used the Swedish National Quality Register for Oesophageal and Gastric Cancer (NREV) to perform traditional multivariable logistic regression and XAI. The 90-day mortality was 6.0%, while anastomosis leakage was present in 12.4%. The XAI models yielded an area under the curve (AUC) of 0.91 for 90-day mortality (as compared with 0.84 for logistic regression). For anastomosis leakage, the AUC was 0.84 using XAI (0.74 using logistic regression). We show that age (mortality increases sharply after 55 years) and body mass index (BMI) (lowest mortality for BMI 30 kg/m^2^) are important survival factors. Additionally, we show that surgery time (minimum anastomosis leakage for a surgery time of 200 min to sharply increase to a maximum at 375 min) and BMI (the lower the BMI, the less anastomosis leakage) are important factors for anastomosis leakage. The surgical understanding of anastomosis leakage and mortality after oesophagectomy is advanced by judiciously applying XAI to structured data. Our nationwide oesophagectomy data contains significant nonlinear relationships. With the help of XAI, we extract personalised knowledge, bringing oesophagus surgery one step closer to personalised medicine.

## Introduction

Oesophagectomy is often performed as a curative measure for oesophageal cancer. It is an extensive surgical procedure. The surgical techniques have evolved, and most of the procedures performed today are more or less minimally invasive. Anastomotic techniques have also developed, and most anastomoses nowadays are performed with circular or linear stapling devices. Perioperative care has also dramatically changed with enhanced recovery programs [1, 2, 3, 4].

Despite these improvements, oesophagectomy still carries a high rate of anastomosis leakage (AL) of 10-15% and a 90-day mortality (D90) rate of 2-5%. AL can be a severe complication that may need complex interventions and increase the risk for D90. To improve the outcome of oesophagectomy, it is necessary to understand which factors affect AL and D90. Modifiable factors are especially important in this context. However, age and other patient-specific features are also important in understanding which patients should or should not be offered oesophagectomy.

Traditional logistic regression models are well-suited for simple relationships and are widely used for predictions in medicine. Basic assumptions that must be met for logistic regression include independence of errors, linearity in the logit for continuous variables, absence of multicollinearity, and lack of strongly influential outliers. As artificial intelligence (AI) continues to integrate into various sectors of society, including advancements in medical research, the pros of more refined prediction models for mortality and anastomosis leakage have become increasingly promising. XGBoost is a recent tool in the AI toolbox that has won several international awards. Shapley scores provide a new way of presenting results, and the two methods combined offer flexibility, predictive performance, interpretability for handling nonlinear relationships, complex interactions, and great visualisations in scientific settings. Together, XGBoost and Shapley scores offer what can be called explainable artificial intelligence (XAI). This study aims to improve the prediction of 90-day mortality and anastomosis leakage after oesophagectomy in a Swedish cohort by using XAI with XGBoost and Shapley scores.

## Methods

### Patients

Data was collected from the National Quality Register for Esophageal and Gastric Cancer in Sweden (NREV). Survival data is automatically transferred to NREV from Statistics Sweden. NREV is well-described, researched and validated [5, 6].

Patients with oesophageal cancer were selected between November 2005 and February 2018, and of these, 1846 patients underwent oesophagectomy for oesophageal cancer. One hundred forty perioperative variables not directly linked with the outcome were selected [7]. All data was extracted on the 11th of March 2020. The study was approved by the Regional Ethical Board of Stockholm (Dnr 2013/596–31/3, amendment: 2020-06495).

### Statistics

We used R for all calculations and graphs. Anastomotic leakage rate and 90-day mortality were modelled using traditional logistic regression and XAI. In the analyses, the predicted event was indicated as 1 (death within 90 days or anastomosis leakage). Anastomosis leakage was defined as a full thickness gastrointestinal defect involving esophagus, anastomosis, staple line, or conduit irrespective of presentation or method of identification with required intevention, surgical or drainage (type II & III) [8]. We excluded variables with more than 20% missing values to ensure the robustness of our analyses. In the remaining dataset, we employed multiple imputations by chained equations with the random forest imputation method using the `MICE` (version 3.16.0) package of `R`.

All pre- and perioperative variables were used for the logistic regressions, and backward elimination was performed separately for 90-day mortality and anastomosis leakage, retaining the variables that best explained these models. These remaining variables were then used in the logistic regression model.

For the XAI, we used the eXtreme Gradient Boosting (`XGBoost`) package (version 1.7.5.1). XGBoost uses decision trees through gradient boosting [9]. We divided our dataset into a 90% training set for model training and a 10% test set for evaluation. Several hyperparameters control XGBoost: the number of decision trees, training rounds, and the learning rate. These hyperparameters must be set before training the algorithm since they significantly impact performance [10]. We used cross-validation to select the number of training rounds and grid search to maximise the algorithm’s accuracy. The final hyperparameters were learning rate 0.1, subsample 0.3, colesample bynode 0.3, reg lambda 6, maximum depth 50, evaluation metric AUC, objective binary logistic.

To assess the model accuracy, receiver operating characteristic (ROC) curves were generated, and the area under the receiver operatic characteristic curve (AUC) was calculated. Differences in AUCs were assessed with the test of DeLong et al.[11]. Additionally, for better interpretability of the XGBoost results, we reported them using Shapley Additive exPlanations (SHAP) [12]. Developed by Lundberg and Lee (2017), SHAP provides a unified and theoretically grounded framework for feature importance analysis, whether traditional logistic regression or advanced machine learning. The contribution of each feature is presented as the absolute mean from each SHAP value. In binary prediction, SHAP values equal the log odds in the regression model [13].

## Results

### 90-day mortality

The overall 90-day mortality (D90) was 6 %. The ROC curve for D90 showed improved predictions for the XGBoost model with an AUC of 0.91 compared to 0.84 for the logistic regression model (p-value <0.05) (See Figure 1). In logistic regression, age was associated with an increased risk of D90 (odds ratio (OR) 1.03 (1.0-1.16)) positive lymph nodes (OR 1.07 (1.03-1.11)), and patients categorised as American Society of Anesthesiologists (ASA) grade 3 had an increased risk for mortality (OR 4.20 (2.08 - 8.69), (3.4 % missing data for ASA scores) while increasing body-mass index (BMI) showed a decreased risk (0.92 (0.87-0.98)). Neither bleeding, surgery time, nor investigated lymph nodes showed any significant association with D90 in the logistic regression model (See Table 1).

**Table 1.**
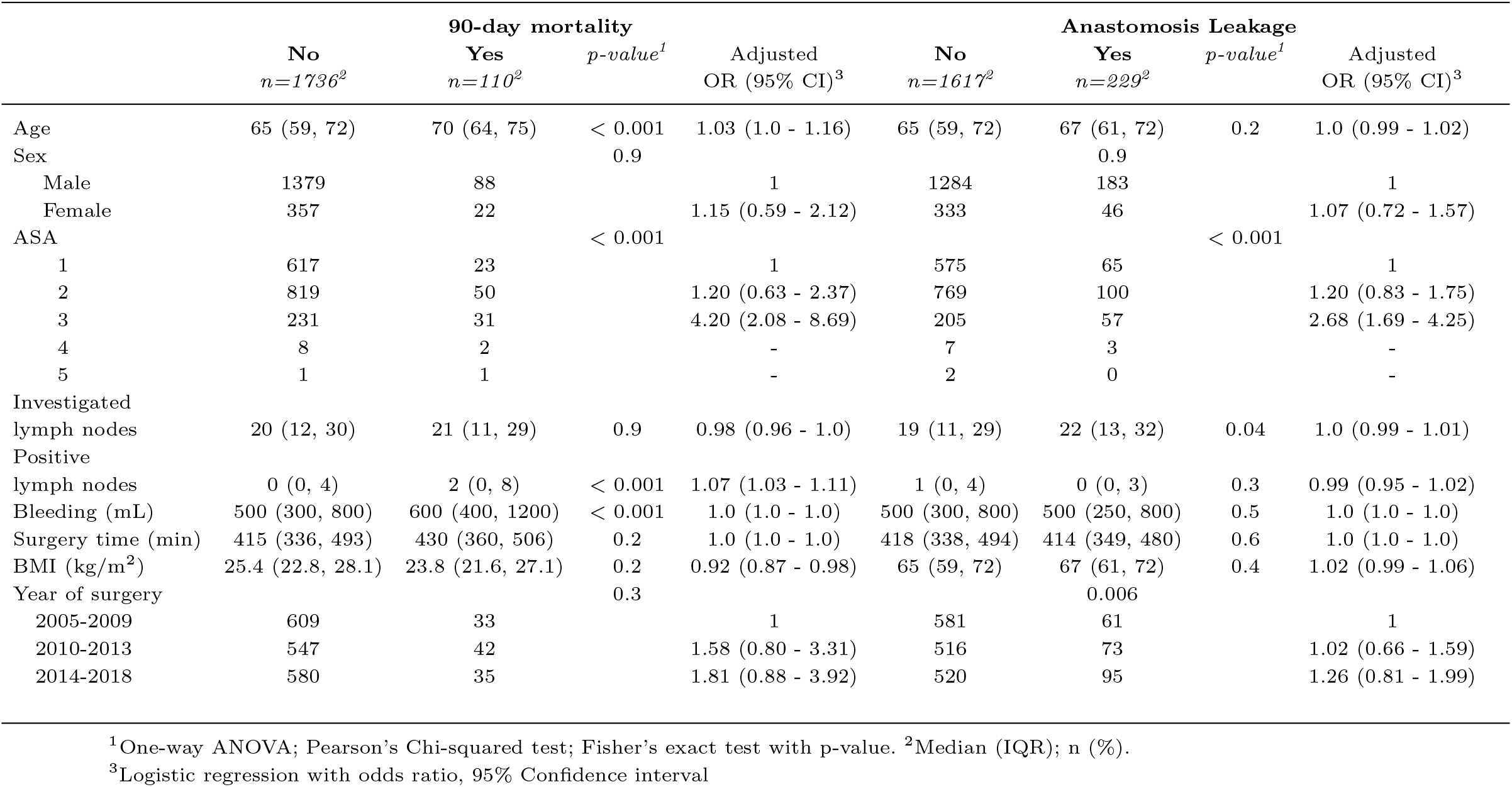
Charachteristics stratified by 90-day mortality and Anastomosis Leakage

**Fig. 1.**
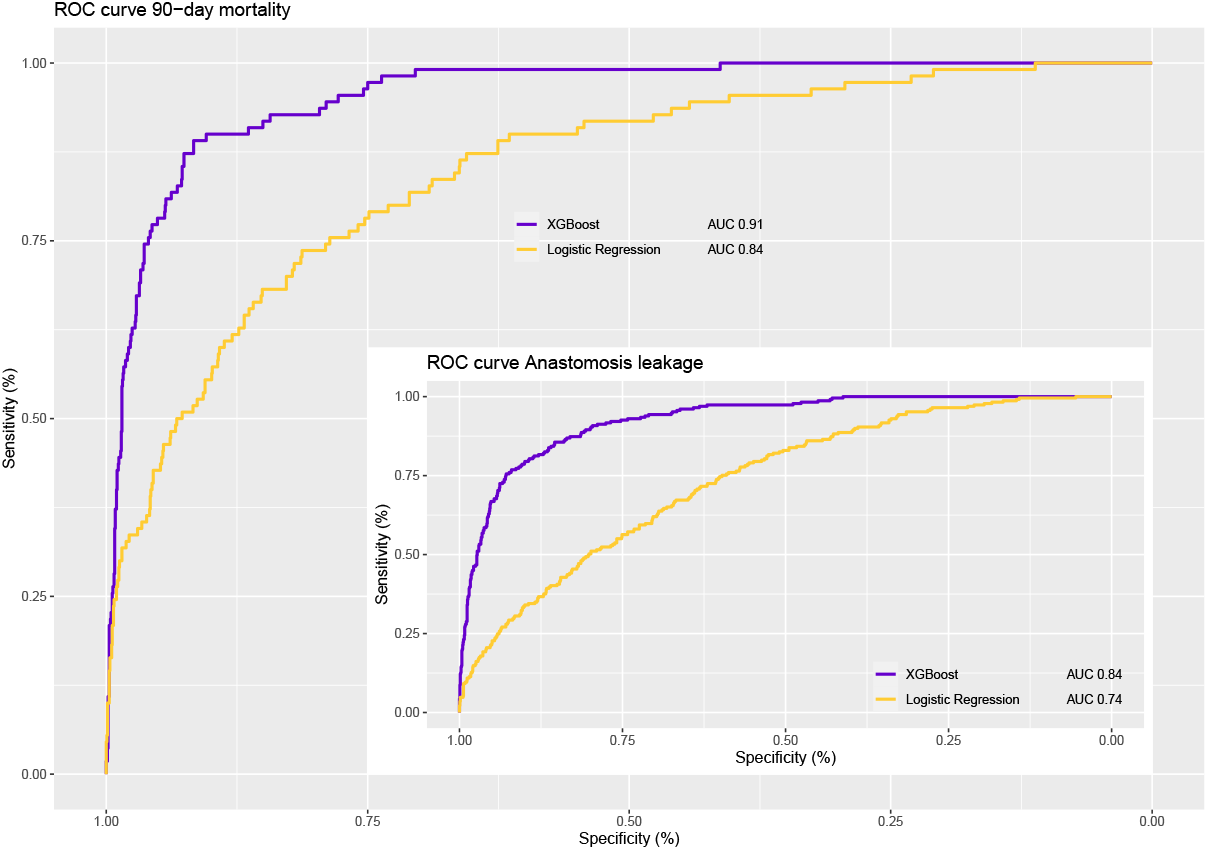
Receiver operatic characteristic (ROC) curves. Main: ROC curve 90-day mortality. Inserted: ROC curve Anastomosis leakage

The XAI analysis found that the most important feature predicting D90 was age (absolute mean 0.36), followed by BMI and bleeding (Absolute mean 0.28, respectively 0.23) (see Figures 2, and 5). Higher age predicted D90, while higher BMI was protective. Females generally had a lower BMI than males. Increased surgery time, the number of positive lymph nodes, and higher ASA grade were also associated with a greater risk of D90. BMI was protective mainly among ASA 1 and 2 while increasing BMI among ASA 3 patients was associated with a greater risk of D90.

**Fig. 2.**
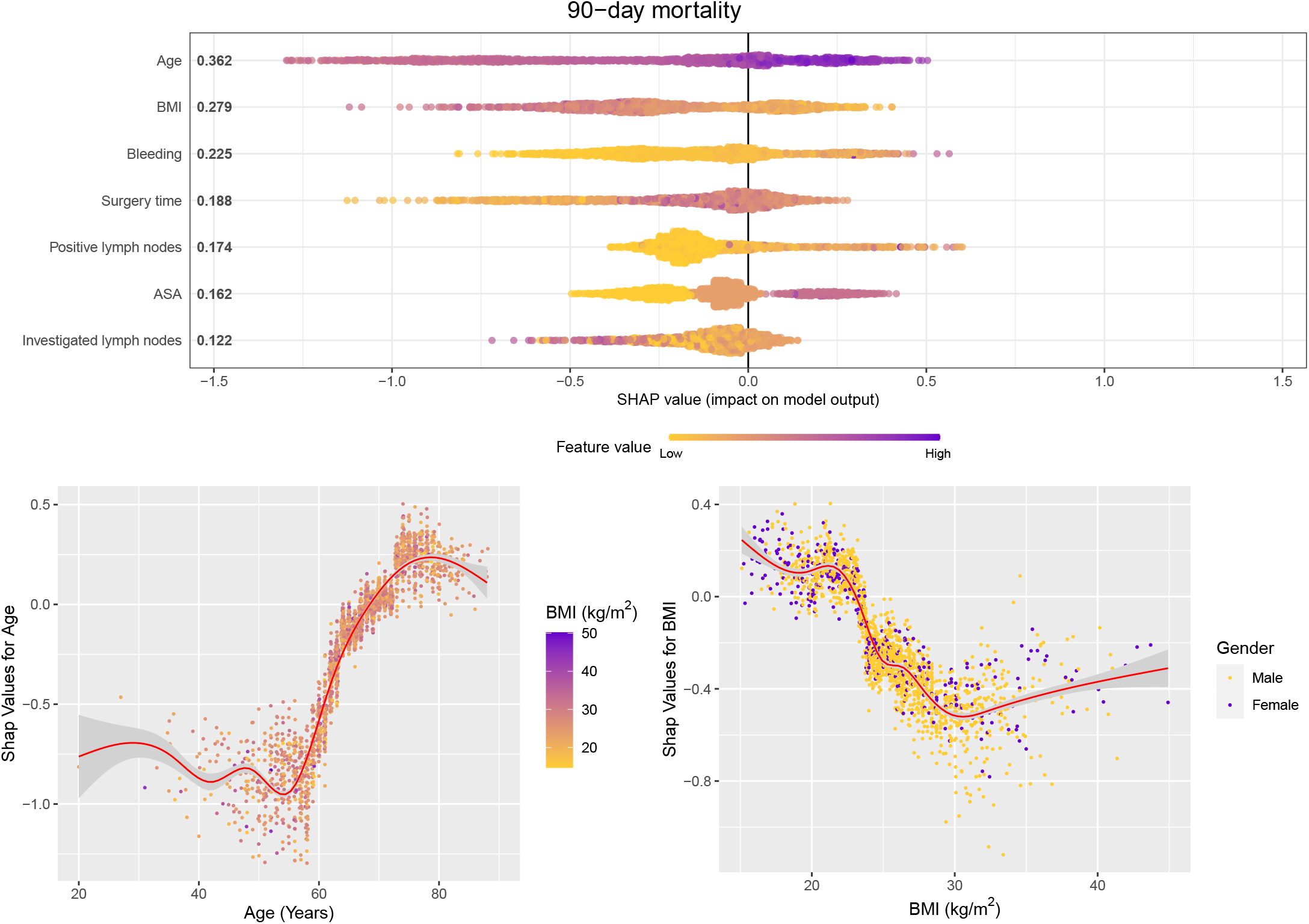
SHAP values for 90-day mortality Top panel: *SHAP Summary plot*. The numbers show the absolute mean Shapley score for each variable, a relative measure of its contribution to predicting D90. The x-axis indicates the SHAP values. Each point in the figure represents a case, and the value on the x-axis indicates the individual SHAP value for that variable. The SHAP value equals the log odds in a regression model. The colour indicates the real value and a darker colour indicates a higher real value (i.e. an age of 80 has a darker colour than 50 years). In summary, age and body mass index (BMI) contributed the most to predicting D90; a higher age increased the risk (darker on the right-hand side), while a higher BMI decreased the risk (brighter on the left-hand side). Bottom panel: *SHAP Dependency plots for Age (left) and BMI (right)*. The points represent cases and show the SHAP value on the y-axis and the real value on the x-axis. The SHAP value equals the log odds in a regression model. The contribution of age to D90 was constantly low for 55 and younger, sharply increasing to a maximum at 75-80 years. The contribution of BMI to D90 decreased with BMI to reach a minimum at 30 kg/m^2^, after which it increased marginally. ASA: *American Society of Anesthesiologist grade*. BMI: *Body mass index*.

**Fig. 3.**
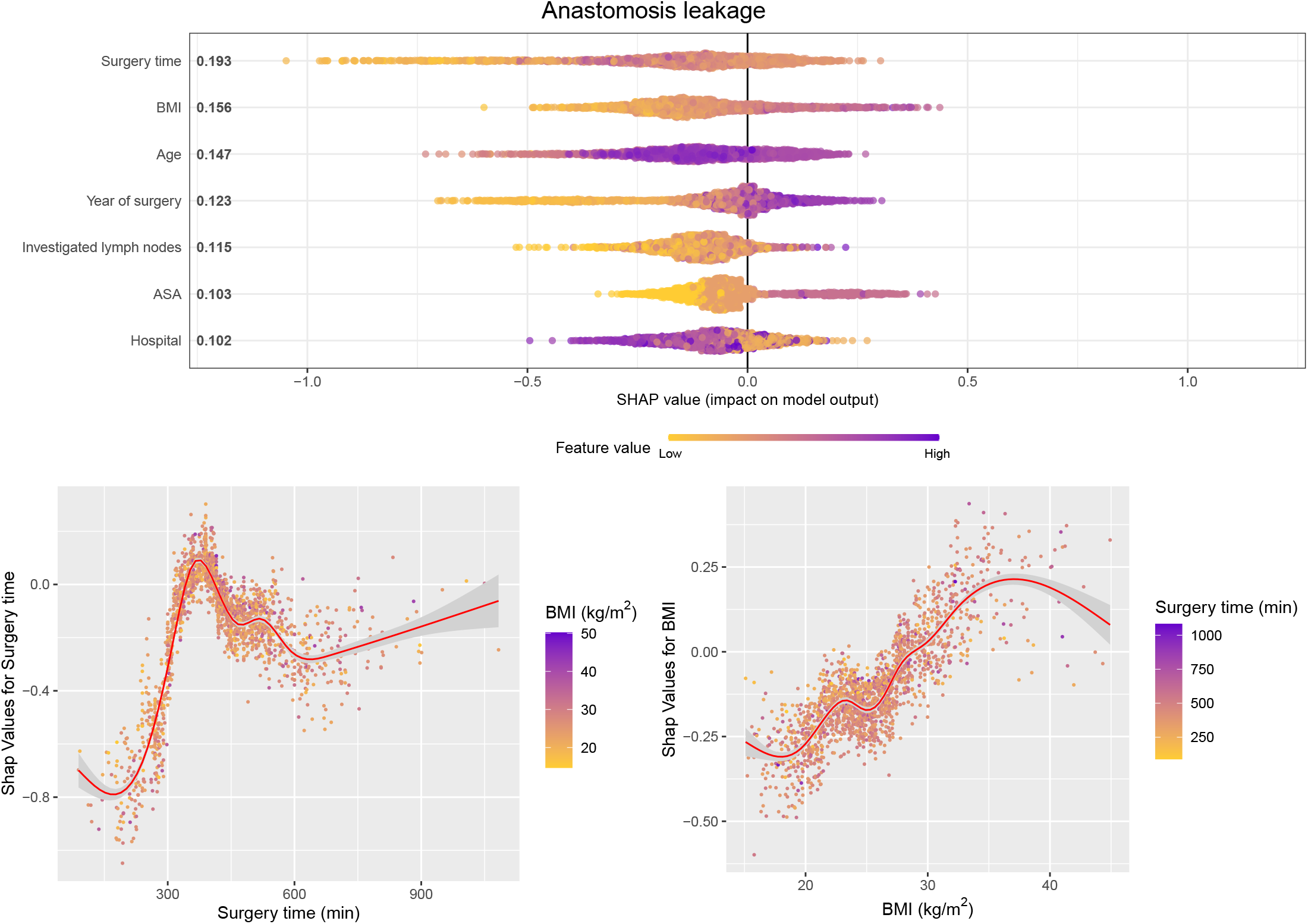
SHAP values for anastomosis leakage Top panel: *SHAP Summary plot*. The numbers show the absolute mean Shapley score for each variable, a relative measure of its contribution to predicting anastomosis leakage. The x-axis indicates the SHAP values. Each point in the figure represents a case, and the value on the x-axis indicates the individual SHAP value for that variable. The SHAP value equals the log odds in a regression model. The colour indicates the real value and a darker colour indicates a higher real value (i.e. an age of 80 has a darker colour than 50 years). In summary, surgery time and BMI contribute the most to predicting anastomosis leakage; surgery times shorter than 300 min decreases the risk (brighter on the left-hand side), while a higher BMI increases the risk (darker on the right-hand side). Bottom panel: *SHAP Dependency plots for Surgery time (left) and BMI (right)*. The points represent cases and show the SHAP value on the y-axis and the real value on the x-axis. The SHAP value equals the log odds in a regression model. The contribution of surgery time to anastomosis leakage increases sharply from a minimum at 200 minutes to a maximum at 375 minutes. The contribution of BMI to anastomosis leakage increases with BMI. ASA: *American Society of Anesthesiologist grade*. BMI: *Body mass index*.

**Fig. 4.**
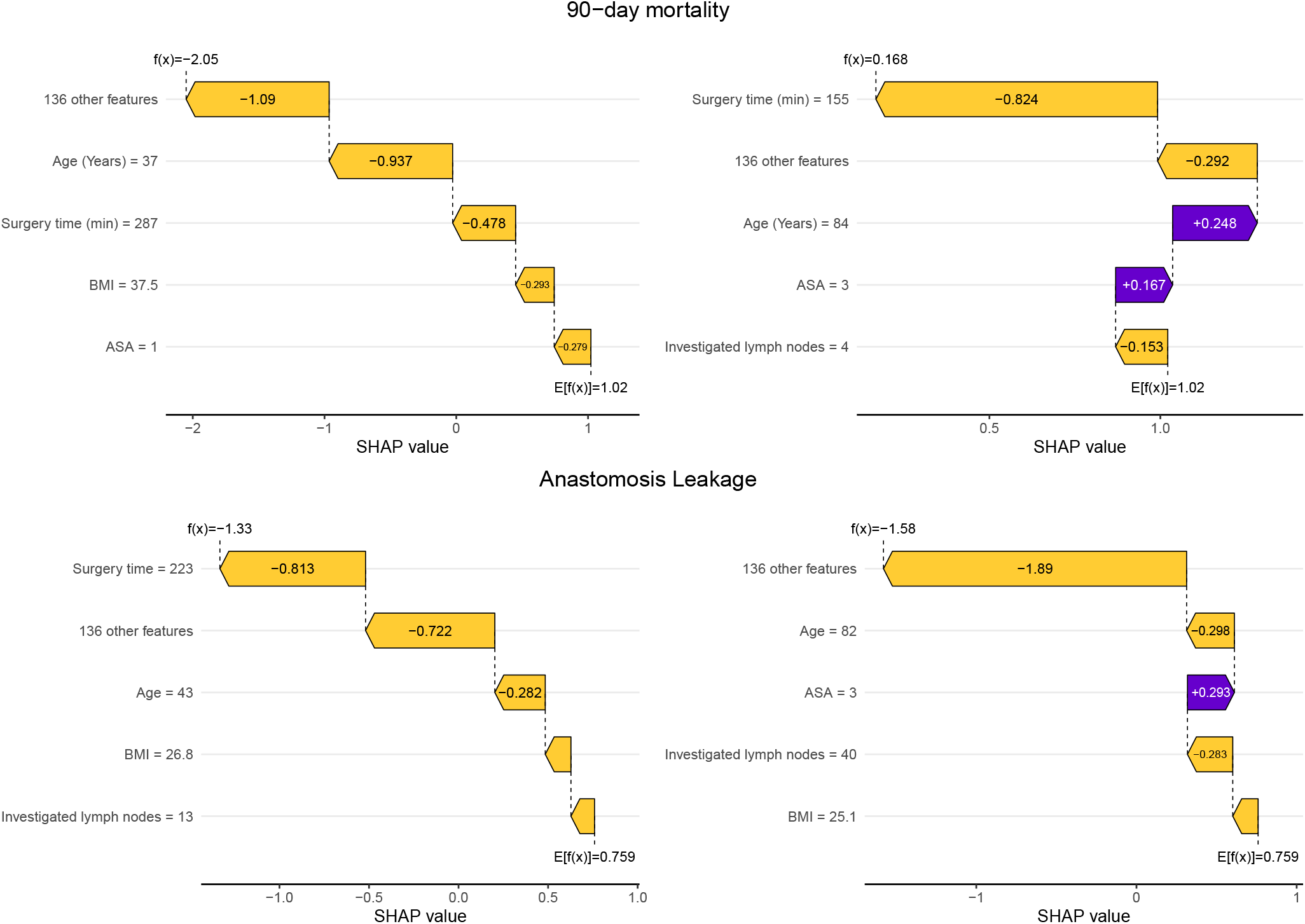
Waterfall plots with SHAP values. The waterfall plots illustrate the additive nature of the Shapley scores in arriving at individual log odds, f(x) for case x, by adding Shapley scores for each feature to the expected log odds for the cohort, E(f(x)). *Top left panel:* The individual log odds for 90-day mortality was -2.05, the sum of the log odds (in decreasing importance) of 136 various features, age, surgery time, body-mass index (BMI), American Society of Anesthesiologist grade (ASA), and the expected cohort value was 1.02. For this individual, the most important contributors to the odds of -2.05 were 136 various features and age (with a negative contribution of -0.937, i.e. decreased risk due to the patient’s age of 37 years). *Top right panel:* The individual log odds for 90-day mortality was 0.168, which is the sum of the log odds (in decreasing importance) of surgery time, 136 various features, age, ASA, investigated lymph nodes, and the expected value of the cohort 1.02. For this individual, the most important contributors to the odds of 0.168 were a short surgery time of 155 min, contributing negatively with -0.824 and 136 various features. *Bottom left panel:* The individual log odds for anastomosis leakage was -1.33, the sum of the log odds (in decreasing importance) of surgery time 223 min, 136 various features, age 43 years, BMI 26.8 kg/m^2^, and 13 investigated lymph nodes, and the expected cohort value 0.759. For this individual, the most important contributors to the odds of -1.33 were a short surgery time of 223 min and 136 various features. *Bottom right panel:* The individual log odds for anastomosis leakage was -1.58, which is the sum of the log odds (in decreasing importance) of 136 various features, age 82 years, ASA grade 3, 40 investigated lymph nodes, BMI 25.1 kg/m^2^, and the expected value of the cohort 0.759. For this individual, the most important contributors to the odds of -1.58 were 136 various features and 82 years old. ASA: *American Society of Anesthesiologist grade*. BMI: *Body mass index*.

**Fig. 5.**
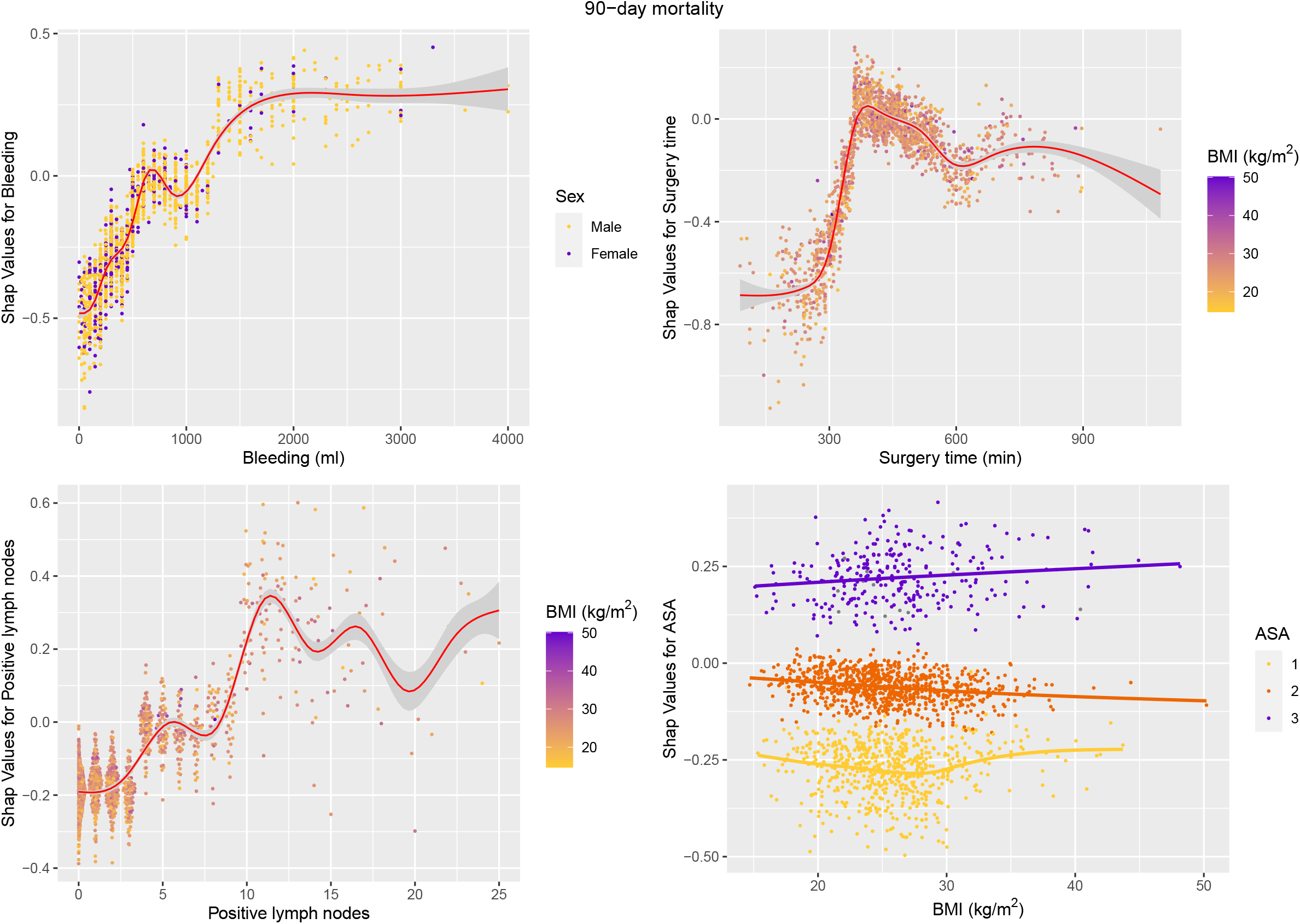
SHAP values for 90-day mortality *SHAP Dependency plots for Bleeding (top left), Surgery time (top right), No. of positive lymph nodes (bottom left) and body mass index (BMI) (bottom right)*. The points represent cases and show the SHAP value on the y-axis and the real value on the x-axis. The SHAP value equals the log odds in a regression model. The contribution of bleeding to D90 increased to reach a plateau for over 1500 mL. The contribution of surgery time to D90 increased sharply from 300 min to a maximum at 400 min, after which it slowly decreased. The contribution of the number of positive lymph nodes increased with the number of lymph nodes to reach a maximum from 11 positive lymph nodes. The contribution of the number of positive lymph nodes increased with the number of lymph nodes to reach a maximum from 11 positive lymph nodes. The contribution of ASA to D90 increased with increasing ASA grade. There was an interaction with BMI. BMI: *Body mass index*.

The contribution of different factors to D90 (and AL) are shown for some example patients in Figure 4.

### Anastomosis leakage

Altogether, 229 patients (12.4 %) developed anastomosis leakage (AL). The AUC for the XGBoost model was 0.84, compared to the AUC of the logistic regression model, which was 0.74 (p-value <0.05) (See Figure 1). In the logistic regression model, ASA 3 patients had a significantly increased risk of developing AL (OR 2.88 (1.69 - 4.25)). No other significant feature was associated with AL (See Table 1).

The XAI analysis results are presented in Figures 3 and 6. The most important feature predicting AL was surgery time (absolute mean 0.19), where the risk for AL peaked around 400 minutes and again declined after that (Figure 3). Higher BMI and an increasing number of investigated lymph nodes were associated with an increased risk of AL (absolute mean 0.16, respectively 0.12). Increased age, the number of investigated lymph nodes, and higher ASA grade were also associated with a greater risk of D90. However, the relationship was not linear, and the risk of AL peaked at around seventy years of age and declined after that. The association with investigated lymph nodes was not linear (see Figure 6).

**Fig. 6.**
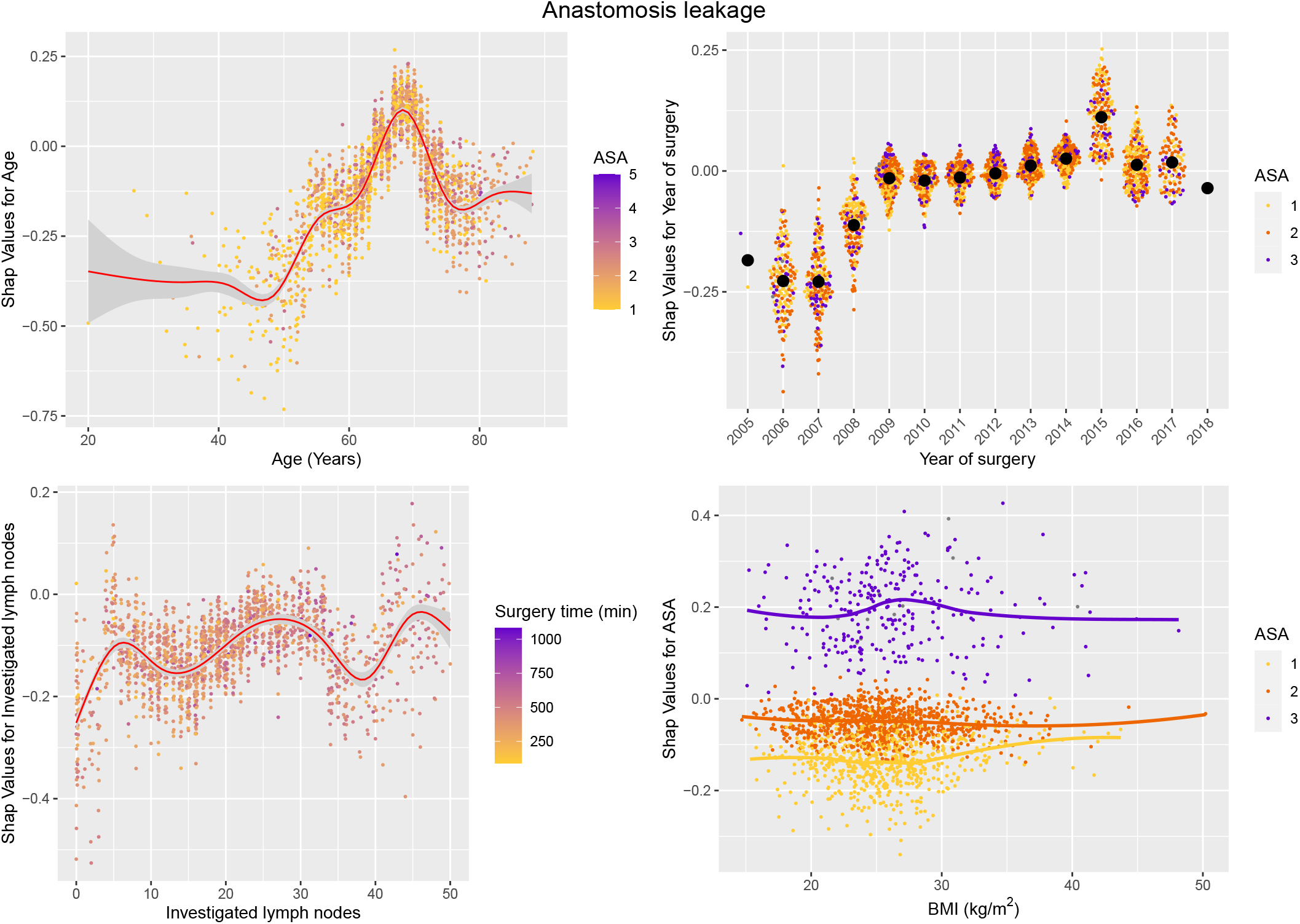
Dependency plots for anastomosis leakage. The contribution of age to anastomosis leakage is shown in the top left panel, displaying a sharp increase of risk from 50 to a maximum risk at 68 years old, after which a limited decline was seen. There was an increased risk for anastomosis leakage over the years, with a sharp increase from 2007 to 2008. Increasing ASA grade carried an increased risk for anastomosis leakage.

The contribution of different factors to AL (and D90) are shown for some example patients in Figure 4.

## Discussion

In this retrospective register-based study, we analysed whether XAI can bring further knowledge in predicting D90 and AL. We have shown that our XAI models are considerably better than our traditional logistic regression models. Further, our model is also better than previous D90 prediction models [14]. Our prediction was better for D90 than AL, although AL had a reasonable AUC. This implies that AL can have other more influential risk factors not accounted for by our variables. Previous medical studies have also shown improved AI prediction models compared to logistic regression models [15, 16].

To elucidate the importance of individual factors on D90 and AL, we decided to employ Shapley scores, a game theoretic approach that can be used with any model, be it AI or logistic, to introduce interpretability. To illustrate the additive contribution of Shapley scores, we have included example Waterfall plots (See Figure 4). This figure exemplifies the relative importance of different factors to D90 and AL for two patients. The choice of XGBoost among many available AI models rests on its proven track record in AI competitions for structured data such as ours.

The data source of this study comprises 100 % of the Swedish operating hospitals for oesophageal cancer and has a more than 95 % coverage rate for the surveys. The completeness and correctness of the surveys are high [5]. A validation program to optimise the completeness and correctness of NREV data, including site visits, has been ongoing for a couple of years.

### 90-day mortality

In previous studies, age and comorbidities have been associated with increased risk of D90 [14, 17], but the XAI demonstrates that age, BMI, bleeding, surgery time, no. of positive lymph nodes and ASA are the most important factors (in falling order) for D90 (see Figure 2).

The XAI highlights how mortality is low for patients less than 55 years old and increases to a maximum for over 75 rapidly. In the logistic regression analyses, BMI exhibited a reduced risk for D90 but was not significant for AL.

Our XAI models show a strong contribution of BMI for both D90 and AL (absolute mean Shapley scores 0.28, respectively 0.16). The Shapley scores show a decreased risk for D90 with a higher BMI (to reach a minimum at BMI 30 kg/m^2^). This aligns with a previous meta-analysis by Mengardo et al. [18]. However, further analysis of D90 and BMI shows an interaction with ASA insofar as higher BMI is associated with lower D90 for ASA 2 and ASA 3 patients, whereas increased BMI for ASA 1 is associated with higher D90.

### Anastomosis leakage

In previous studies, comorbidities and BMI have been associated with increased risk of AL [18], but the XAI demonstrates that surgery time, BMI, age, year of surgery, investigated lymph nodes, and ASA were the most important factors (in falling order) for AL (see Figure 3).

Increasing surgery time is the most important factor for AL. It is lowest for 200 min to rapidly reach a maximum for 350 min to again decrease to a lesser degree until 600 min surgery duration.

The logistic regression analyses BMI showed no significant association for AL, but the Shapley scores for the XAI method showed a high contribution to the model and an increased risk of AL with increasing BMI (to reach a maximum at BMI 35 kg/m^2^), in line with a Mengardo et al. [18].

Since participation in the NREV quality register is not mandatory but highly recommended, results from the first few years may carry some bias. An interesting example of this phenomenon was that the risk for anastomosis leakage increased dramatically from low reported values between 2005 and 2008, and later on, it still increased but at a much slower rate. A transiently high risk was seen in 2015. The slow increase in risk during the last 10 reported years could be attributed to the fact that the surgical centres have become more active in identifying anastomosis leakages. Enhanced Recovery Programmes (ERP) were introduced with new postoperative routines. Endoscopic and improved radiological evaluations were introduced more liberally, and some previously missed subclinical leaks were probably reported. The introduction of laparoscopic and robotic techniques during the study period might impact the increased frequency of anastomotic leakage.

The significantly better predictions with the XAI method result from complicated nonlinear dependencies of the covariates, validating the use of a much more complex and less intuitive model than classical multivariable logistic regression. An illustrative instance of this phenomenon is BMI, which holds significant predictive value for D90 and AL. Notably, a BMI of 30 emerges as optimal for reducing the risk of D90, with lower and higher BMI values correlating with increased risk (see Figure 2).

A strength of our study is that we employ a high-quality national registry with a high coverage rate and few missing data points [5]. Previous NREV studies on long-term outcomes after oesophagectomy showed the impact of sex, education level, and geographical differences within the country [6, 19, 20]. In this study, with early postoperative follow-up, we evaluated the impact of sex and, to some extent, geographical difference by adjustments for operating hospitals, factors in this study that were of limited impact.

A weakness of this registry is that, for some reason, it has low coverage of tobacco use, which, therefore, was excluded from the analysis. The impact of detailed oncological therapy was also limited in this study. This was compensated by the fact that patients who underwent oesophagectomy during the study period all had perioperative oncological treatment according to international recommendations for their tumour stages.

A possible extension to the machine learning methodology would be to develop formal testing methods for Shapley scores, which is beyond the scope of the present paper.

## Conclusion

To summarise, we have advanced the knowledge of risk factors for 90-day mortality and anastomosis leakage after oesophagectomy by using explainable AI (XAI). To mention the main findings, we show that age (mortality increases sharply after 55 years) and BMI (lowest mortality for BMI 30 kg/m^2^) are important survival factors. Additionally, we show that surgery time (minimum anastomosis leakage for a surgery time of 200 min to sharply increase to a maximum at 375 min) and BMI (the lower the BMI, the less anastomosis leakage) are important factors for anastomosis leakage. In a more general sense, we advance the surgical understanding of anastomosis leakage and mortality after oesophagectomy by judiciously applying XAI to structured data. Our nationwide oesophagectomy data contains significant nonlinear relationships. With the help of XAI, we extract personalised knowledge, bringing oesophagus surgery one step closer to personalised medicine.

## Data Availability

All data produced in the present work are contained in the manuscript

## Author contributions statement

JJ conceived and funded the study. SD conducted all the calculations advised by AF. SD and AF wrote the first draft of the manuscript. All authors read and revised the final manuscript.

## Supplement

